# Novel intraoperative online functional mapping of somatosensory finger representations for targeted stimulating electrode placement

**DOI:** 10.1101/2020.07.10.20149021

**Authors:** David P. McMullen, Tessy M. Thomas, Matthew S. Fifer, Daniel N. Candrea, Francesco V. Tenore, Robert W. Nickl, Eric A. Pohlmeyer, Christopher Coogan, Luke E. Osborn, Adam Schiavi, Teresa Wojtasiewicz, Chad Gordon, Adam B. Cohen, Nick F. Ramsey, Wouter Schellekens, Sliman J. Bensmaia, Gabriela L. Cantarero, Pablo A. Celnik, Brock A. Wester, William S. Anderson, Nathan E. Crone

**Affiliations:** National Institute of Mental Health, National Institutes of Health, Bethesda, Maryland, USA; Department of Biomedical Engineering, Johns Hopkins University, Baltimore, Maryland, USA; Research and Exploratory Development Department, Johns Hopkins University Applied Physics Laboratory, Laurel, Maryland, USA; Department of Physical Medicine and Rehabilitation, Johns Hopkins University, Baltimore, Maryland, USA; Department of Neurology, Johns Hopkins University, Baltimore, Maryland, USA; Department of Anesthesiology and Critical Care Medicine, Johns Hopkins University, Baltimore, Maryland, USA; Department of Neurosurgery, Johns Hopkins University, Baltimore, Maryland, USA; Department of Plastic and Reconstructive Surgery, Johns Hopkins University, Baltimore, Maryland, USA; UMC Utrecht Brain Center, Utrecht, Netherlands; Department of Organismal Biology and Anatomy, University of Chicago, Chicago, IL, 60637, USA

**Keywords:** intraoperative functional mapping, online functional mapping, brain-computer interface, brain-machine interface, intra-cortical microstimulation, microelectrode array (MEA), electrocorticography (ECoG)

## Abstract

Defining eloquent cortex intraoperatively, traditionally performed by neurosurgeons to preserve patient function, can now help target electrode implantation for restoring function. Brain-machine interfaces (BMIs) have the potential to restore upper-limb motor control to paralyzed patients but require accurate placement of recording and stimulating electrodes to enable functional control of a prosthetic limb. Beyond motor decoding from recording arrays, precise placement of stimulating electrodes in cortical areas associated with finger and fingertip sensations allows for the delivery of sensory feedback that could improve dexterous control of prosthetic hands. In our study, we demonstrated the use of a novel intraoperative online functional mapping (OFM) technique with high-density electrocorticography (ECoG) to localize finger representations in human primary somatosensory cortex. In conjunction with traditional pre- and intraoperative targeting approaches, this technique enabled accurate implantation of stimulating microelectrodes, which was confirmed by post-implantation intra-cortical stimulation of finger and fingertip sensations. This work demonstrates the utility of intraoperative OFM and will inform future studies of closed-loop BMIs in humans.

## Introduction

Functional localization has long been utilized by neurosurgeons primarily to spare eloquent cortex during resection surgeries.^1–5^ However, novel neurotechnologies can utilize functional localization for targeting of restorative neural implants. Brain-machine interfaces (BMIs) have the potential to restore function to paralyzed patients via brain control of neuroprostheses.^6–11^ Building on evidence demonstrating the importance of finger and fingertip sensations in upper-limb control^12^, sensory feedback through cortical stimulation has been incorporated into closed-loop BMIs to improve motor control.^13^ To guide implant placement within hand or arm representations in somatosensory cortex, BMI researchers have traditionally relied on preoperative neuroimaging in humans^14,15^ or surgical atlases in non-human primates (NHP).^16^ However, the development of fully dexterous neuroprostheses requires novel surgical approaches to precisely place stimulating electrode arrays in finger eloquent cortex.

Online functional mapping (OFM) can provide high temporal and spatial information to clinicians in real-time to assist in functional localization. OFM displays task-based cortical activity gathered from electrocorticography (ECoG) electrodes placed on the surface of the brain. It has been used to define eloquent cortex in the epilepsy monitoring unit^17–20^ and intraoperatively.^21,22^ For closed-loop BMIs, a high-density ECoG (hd-ECoG) array can maximize the opportunity of identifying finger-specific sensory activations at a high spatial resolution.^23,24^ Combining OFM with more traditional preoperative (neuroimaging) and intraoperative (neuromonitoring, neuronavigation) approaches can provide a more defined map of finger representations in somatosensory cortex.

Here we report the novel use of hd-ECoG intraoperative OFM during vibrotactile stimulation of individual fingers to guide placement of stimulating microelectrode arrays (MEAs) within finger and fingertip regions in bilateral somatosensory cortices in a paralyzed patient (Fig. 1). We present preoperative and intraoperative strategies for optimizing sensory electrode placements with confirmation via post-operative intracortical microstimulation (ICMS).

**FIG. 1.**
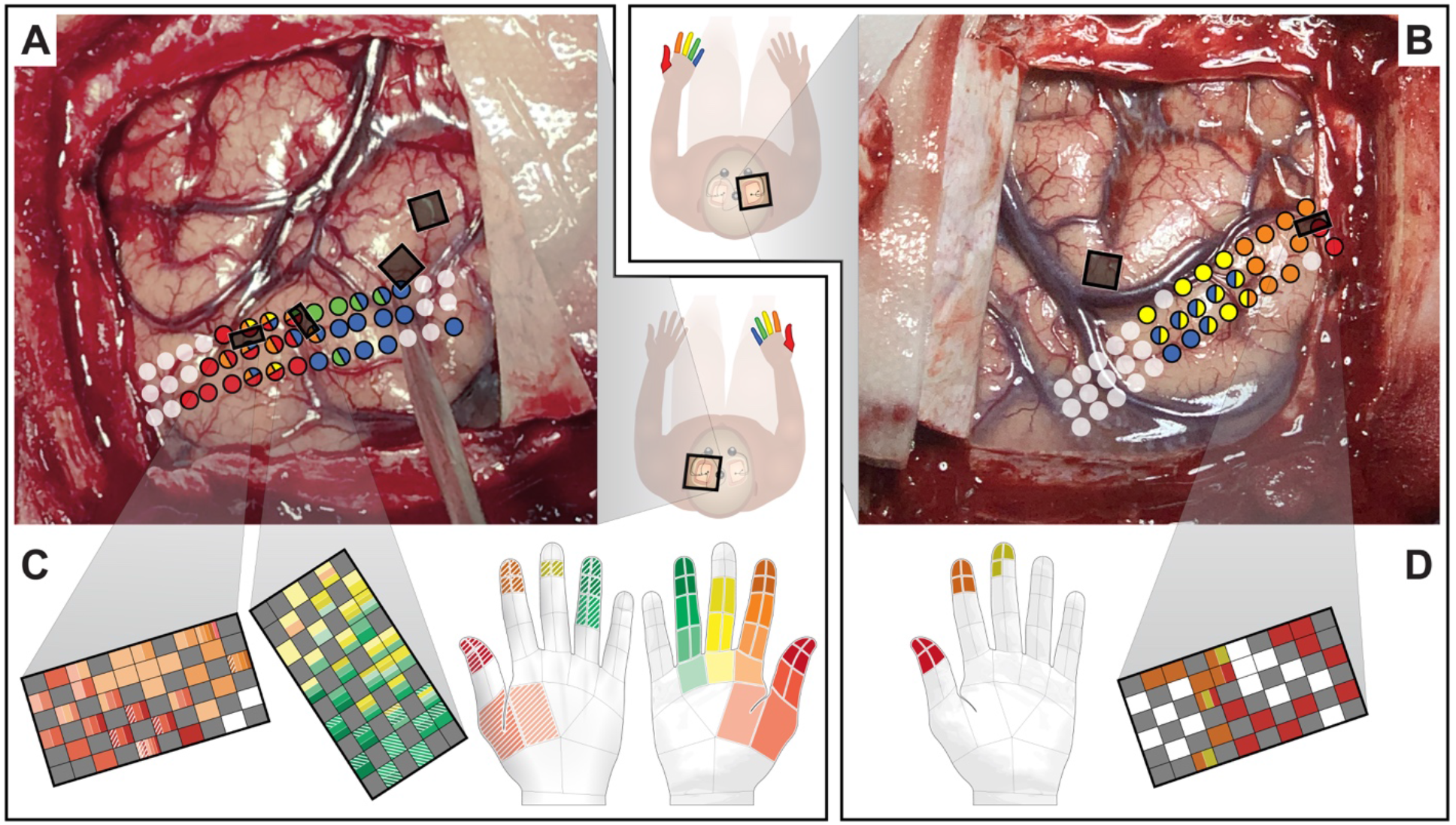
Summary overview of the intraoperative OFM mapping process. High density ECoG grids were placed on the pre- and postcentral gyrus for mapping purposes (only the postcentral gyrus grids are shown for clarity). Finger sensory stimulation was provided via vibrotactile motors to the right (**A**) and left (**B**) hands with resultant high gamma results mapped onto the ECoG grid (grid colors correspond to finger-specific responses). Black boxes on the cortex demarcate final implant locations as determined by post-implantation photographs. Post-operative intra-cortical microstimulation results are shown for the left (**C**) and right (**D**) hemisphere arrays. Projected fields are marked on the MEA and hands with corresponding colors (that is, stimulation at the specific electrode elicited sensations at finger regions indicated by the colors). Solid colors indicate the palmar surface of the hand while hashed colors indicate the dorsal surface.

## Methods

### Patient History

A man in his 40’s with C5-C6 ASIA B tetraplegia (several decades post-injury) gave informed consent for a chronic bilateral implant study under a protocol approved by the Johns Hopkins Institutional Review Board, NIWC Pacific IRB, and FDA IDE 170010. Clinical assessments showed intact sensation to light touch stimulation on all fingers, but only partial sensation to pinprick (intact on first two digits and absent on last three digits of each hand).

### Preoperative Planning

Two months prior to surgery, structural and functional neuroimaging (fMRI) were obtained. For motor mapping, the patient was instructed to attempt and imagine various movements with each upper limb. For sensory mapping, a wire assembly was used to deliver peripheral sensory stimulation by pulling on glove-fitted rings in sync with videos of stimulation on individual fingers (thumb, index, pinky) on each hand. Preoperative motor and sensory implant locations were determined utilizing 3-dimensional reconstructed functional maps (Fig. 2A). Sensory implant sites targeted right- and left-hand thumb/index and right-hand ring/pinky activations.

**FIG. 2.**
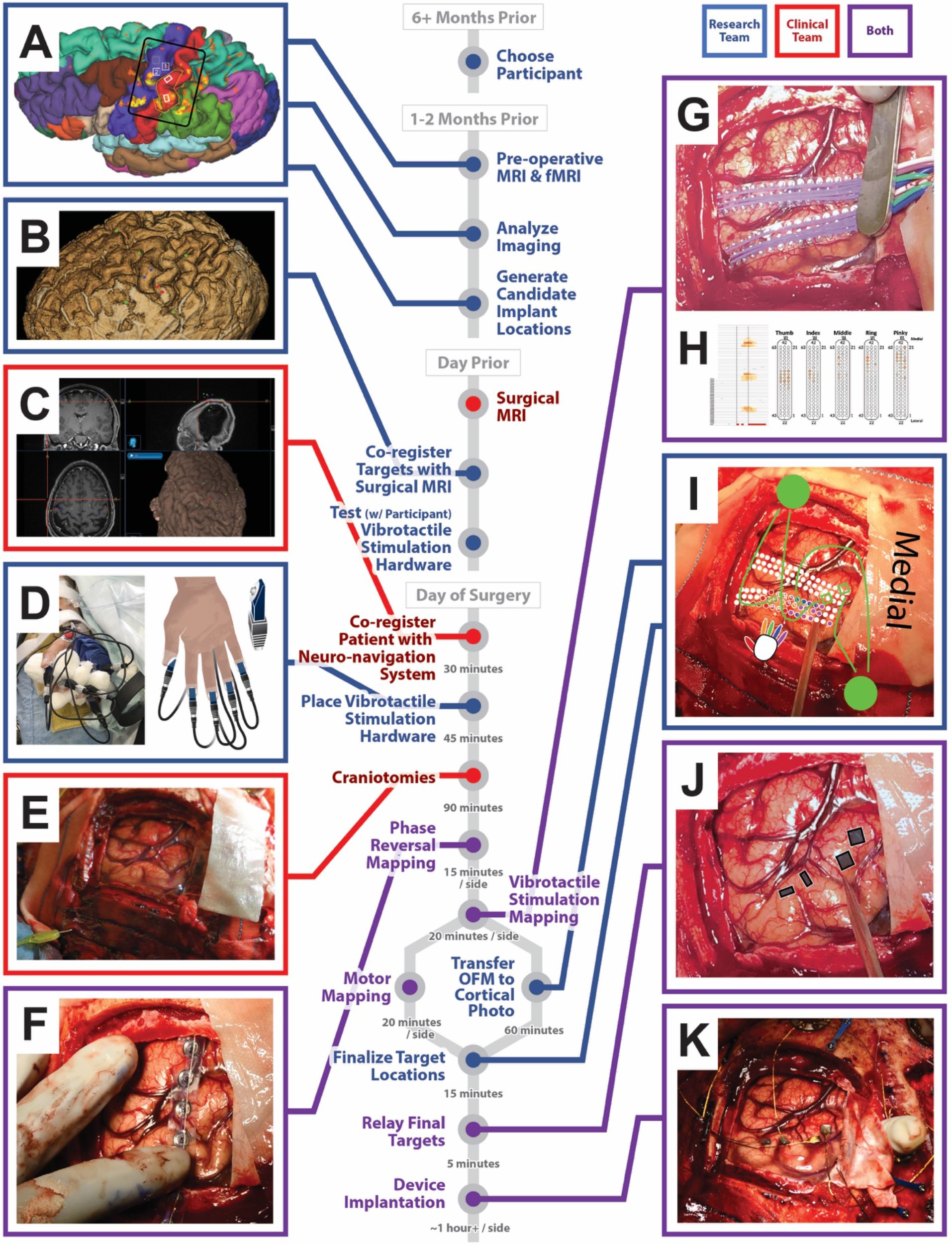
Flow chart of stimulating electrode placement timeline. A patient is recruited six or more months prior to surgery. One to two months prior to surgery, the patient undergoes pre-operative MRI and fMRI imaging which is analyzed to generate a set of candidate implant locations (A). The day before surgery, the patient undergoes a surgical MRI for use in the neuronavigation system and candidate implant sites are co-registered with the MRI (B). The vibrotactile stimulators are tested with the patient the day prior to surgery to ensure finger-specific sensation. The day of the surgery, the patient MRI is co-registered with the patient through the neuronavigation system (C) to provide real time feedback of candidate implant targets. The vibrotactile stimulation hardware is placed prior to incision (D). After the craniotomies (E), a 1×8 ECoG strip is placed for phase reversal mapping of central sulcus (F). The novel intraoperative mapping, utilizing high density ECoG arrays (G), is performed to refine finger-specific implant targeting. High gamma activity of vibrotactile stimulation of individual fingers is mapped onto ECoG figures (H). These results are combined and transferred manually to a diagram overlaid on intraoperative photographs (I) so that the team can debate final target locations and pedestal/wire arrangements (I, in green). The final targets locations (J) are relayed to the surgeon via a tablet interface for device implantation (K).

The day before the surgery, the patient underwent a high-resolution surgical MRI scan to be utilized in the neuronavigation system. The preoperative implant targets were manually co-registered to the surgical MRI scan (Fig. 2B). Co-registration of the fMRI results failed due to technical considerations. The patient was also introduced to vibrotactile stimulators to assess perceptual quality. The intensity and frequency of vibration was optimized to isolate sensations to individual fingers.

### Surgical and Anesthesia Protocol

The patient underwent an asleep-awake-asleep surgery protocol. The patient was placed in a supine position with both arms slightly abducted to allow access to the palmar finger areas. Induction of anesthesia was performed with 100 mg lidocaine, 200 mg propofol, and was maintained with sevoflurane via a laryngeal mask airway (LMA). After placement of Mayfield pins, the scalp was infiltrated with 20 ml of 0.5 % bupivicaine. A dexmetetomidine infusion at 0.4 mcg/kg/hr was initiated prior to removal of the LMA and emergence from anesthesia and then weaned to optimize awake mapping, which included no additional sedation. Upon completion, sedation with dexmetetomidine 0.4 mcg/kg/hr and propofol 50mcg/kg/min was used until surgical closure. Oro-tracheal intubation was performed with a fiberoptic bronchoscope to facilitate closure and general anesthesia was maintained with sevoflurane.

### Intraoperative Mapping: Traditional Approach

After initial sedation, the patient was co-registered to the Medtronic StealthStation S7 Navigation system (Fig. 2C), which was used to guide craniotomy locations (Fig. 2E) and provide an initial assessment of the preoperative implant targets relative to the underlying cortical anatomy. The central sulcus was localized by performing intraoperative neurophysiologic monitoring (IONM). A 1×8 ECoG strip was placed perpendicular to putative central sulcus, and phase reversal of the evoked potentials was recorded during median nerve stimulation according to established clinical protocol (Fig. 2F).

### Intraoperative Mapping: Novel Online Functional Mapping Addition

The patient was then awakened for intraoperative OFM, performed first on the left hemisphere, followed by the right. In each hemisphere, sensory mapping was followed by motor mapping (each session took about 20 minutes/hemisphere).

Two 3×21 hd-ECoG strips (1 mm contacts, 3 mm spacing) were placed along the central sulcus to cover the pre-central and post-central gyri (Fig. 2G). Broadband high-gamma activity (70-110 Hz), analyzed similar to previous studies,^18,20^ was mapped onto computer renderings of the ECoG strips during vibrotactile stimulation of each finger on the contralateral hand (Fig. 2H). The left ring finger was not mapped because it was not needed for setting or bounding implant targets. Prior to starting the craniotomies, vibrotactile motors were attached to fingertips on both hands with soft cotton dressings used to isolate the fingers from neighboring vibrotactile stimuli (Fig. 2D). Each finger was stimulated 50 times, each at 60Hz for 500ms followed by a jittered 1.5-2.5s pause.

To visualize finger-specific activation patterns on the cortical anatomy, sensory OFM results were manually transferred in real-time, using Adobe Illustrator (Adobe Inc, San Jose, USA), to a grid diagram overlaid on a photograph of the brain surface (Fig. 2I). Similar maps were created while the patient was instructed to attempt hand gestures, but these data were not used to inform motor MEA placement due to inconclusive results (poor patient involvement due to anesthesia).

## Results

### Online Functional Mapping Results

High gamma finger activation patterns (Fig. 2H) were combined and mapped onto an intraoperative photograph to enable comparisons and implant targeting (Fig. 2I). The finger activity followed a clear medial to lateral somatotopic arrangement of pinky to thumb (Fig. 1A/B). Based on these results, the team selected new targets (vs. preoperative targets) for sensory MEA placement, targeting thumb/index cortex and middle/ring cortex in the left hemisphere, and thumb/index cortex in the right hemisphere (Fig. 2I, green arrays).

### Implantation

Final target locations were displayed to the surgeon via tablet interface (Fig. 2J). In total, six MEAs (Blackrock Microsystems, Salt Lake City, Utah) were implanted bilaterally in the patient’s motor and somatosensory cortices (Fig. 2K). Two pairs of arrays were implanted in the left hemisphere, and one pair was implanted in the right hemisphere. Each pair consisted of a 96-channel motor MEA (4×4mm, platinum tip) and a 32-channel sensory MEA (4×2.4mm, sputtered iridium oxide film) wired to a skull-mounted transcutaneous pedestal. Of the four available MEA pairs, three pairs with the least number of channels with impedance outside the acceptable range (> 800 kΩ for motor, > 80 kΩ for sensory MEA) were selected to be implanted into the brain. Arrays were implanted using a high-speed pneumatic inserter.^11^

### Postoperative Confirmation

Structural confirmation consisted of overlaying photographs of the surgical site before and after insertion of implants onto the intraoperative ECoG activation maps to compare the final implant location with preoperative targets derived from fMRI and intraoperative targets derived from OFM (Fig. 3). For the stimulating electrodes, there was an average shift (measured array center to center) of 4.3mm from preoperative to intraoperative targets (4.8mm shift from preoperative to final implant location) with a smaller 2.3mm shift from intraoperative target to final implantation location. A comparison with a postoperative CT scan was uninformative due to significant shifting of the brain within the skull after a lengthy surgery.

**FIG. 3.**
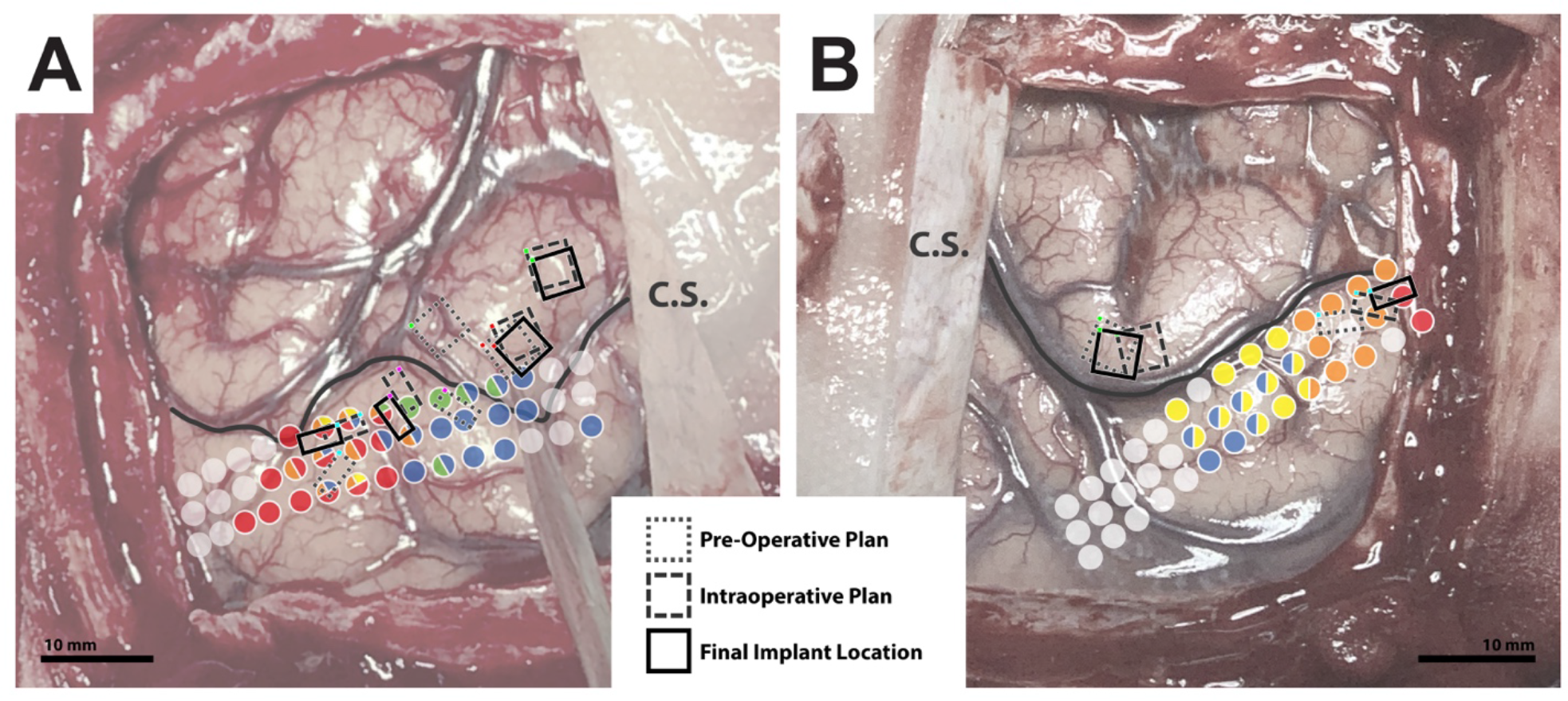
Comparison of pre-operative (dotted lines), intraoperative (dashed lines), and post-operative (solid lines) implant locations for left (A) and right (B) hemisphere. Array implant target sequences are noted by similar color markings in the corner to assist identification of single implants. The arrays are overlaid on intraoperative OFM results to highlight differential functional location of pre-operative and intended stimulation plans. The large shift in the medial left hemisphere motor cortex recording array denoted by a green corner in (A) was due to surface vasculature, which can be seen inside the pre-operative dotted array square. As noted in the text, the pre-operative stimulating electrode planned orientations were already rotated versus the original plan at the time of the surgery. C.S. indicates central sulcus.

The implant locations were also explored functionally by means of ICMS, as described in more depth^25^. In the left hemisphere, stimulation evoked right-hand thumb and index finger and fingertip percepts from the lateral array, and right-hand middle and ring finger and fingertip percepts from the medial array (Fig. 1C). Stimulation of the right hemisphere array evoked left-hand thumb, index, and middle fingertip percepts (Fig. 1D).

## Discussion

We report a novel technique of intraoperative functional mapping to guide placement of stimulating electrode arrays in somatosensory cortex, which enabled finger- and fingertip-specific stimulation in three cortical surface arrays implanted across two hemispheres (Fig. 1). As BMI capabilities advance, there is an increased need to precisely target sensory areas to enable dexterous closed-loop control. Our technique builds upon traditional approaches which rely on preoperative imaging to target implantation of microelectrode arrays. The systematic approach detailed above can be utilized by clinical researchers interested in precisely mapping eloquent cortex for chronic human BMI studies and other surgical purposes.

We utilized a three-axis approach for targeting anatomical fingertip somatosensory representations. The first axis is a rostral-caudal determination of central sulcus to identify the postcentral gyrus. Reconstructions derived from preoperative imaging allowed team members to familiarize themselves with the patient’s unique peri-hand knob gyral anatomy to increase intraoperative surface anatomy recognition (aided by intraoperative neuronavigation). IONM SSEP helped confirm the central sulcus.

The second axis, a medial-lateral sensory somatotopy along postcentral gyrus, was explored to identify finger-specific regions. Preoperatively, we utilized literature studies^3,4,26,27^ and our prior finger functional mapping studies^23^ to determine a priori regions of interest (ROI) spanning from immediate posterior to the hand knob to ∼2 cm lateral along postcentral gyrus.^27,28^ Preoperative fMRI narrowed our ROI, primarily for right thumb and left index, with less information gained for medial ring/pinky activations. Once OFM demonstrated clear somatotopy, it took precedence in determining final targets (Fig. 2I). There was a clear difference between the preoperative and intraoperative plans, with an average MEA shift of 4.3mm.

The third axis, a rostral-caudal finger base to fingertip axis, was determined through OFM of peripheral vibrotactile stimulation of patient fingertips. Prior work demonstrated an intra-digit somatotopy within sensory cortex ^29,30^ and along the crown of the postcentral gyrus.^4^ Our arrays were designed with higher electrode density on the caudal portion to take advantage of this finding (though the orientation of the implants rotated during surgery). However, as we did not peripherally stimulate other regions of the finger (beyond spread of vibrations), it is unclear if our intraoperative approach clearly defined fingertip versus non-fingertip regions or indicated general finger activity. Future studies could clarify these results by stimulating proximal finger areas to contrast with fingertip representations and optimizing peripheral stimulation type (e.g., local electrocutaneous).

### Limitations and Next Steps

The current procedure was conducted with a single patient, which may limit generalizability. However, we successfully demonstrated use of this approach for all three stimulating arrays across two hemispheres, which suggests procedure reproducibility.

Another major question is the generalizability to patients with complete spinal cord injuries (SCI). However, recent studies have demonstrated retained sensory pathways in some patients deemed clinically complete SCI, termed sensory discomplete.^31–33^ Patients with clinically complete injuries could be evaluated prior to surgery for the presence of sensory discomplete lesions (e.g, fMRI activity present for finger sensations), which could benefit from the same approach described in this study.

Implanting patients with truly complete SCI (i.e. minimal/no sensory activation) may entail additional localization difficulties. Preoperative structural imaging can provide historical group ROI coordinates for finger activation (posterior to hand knob and 1-2 centimeters lateral, with fingertips represented caudally), but this may not lead to confidence in targeting individual patient anatomy. Functional neuroimaging of attempted finger movements has the potential to show finger-specific activation patterns, as demonstrated in amputee patients^34^ and a combination of imagined and actual sensory stimulation in another chronic BCI study.^15^ However, targeting surgical implants within areas of neuroimaging activation may be imprecise.^35^ Non-invasive brain stimulation holds potential for functional mapping, with focused ultrasound^36^ demonstrating better focality than transcranial magnetic stimulation^37^ for hand sensory stimulation, but further work is needed to demonstrate reliable fingertip-specific percepts. Intraoperative OFM could potentially be used to define lateral margins of finger ROIs by demarcating facial sensory representations, which would most likely be present immediately lateral to the critical thumb area.^4^

Another approach in complete SCI patients would be to conduct awake electrical cortical stimulation (ECS), which could provide evidence for finger sensory representations.^3,4^ However, eliciting positive reports of sensory percepts instead of negative (numbness) reports or inactivating motor functions transiently^5^ may be difficult,^38^ and prior studies have demonstrated low specificity and variability of cortical stimulation,^39^ including overlapping activations of multiple fingers due to direct cortical stimulation.^40,41^ Recent studies with high density (3mm inter-electrode distance) ECoG-based stimulation have demonstrated higher degree of individual finger specificity,^42,43^ although it is unclear if this approach could delineate fingertip versus more proximal finger/palm regions. Researchers have found a correlation between cortical stimulation, perception of sensation, and high gamma activity, pointing to overlap of the OFM and ECS methods.^44^

A technical area for improvement includes automating display of real-time OFM results. Initially, OFM results were only displayed on grid diagrams (Fig. 2H), but were subsequently added manually via editing software, to the intraoperative photographs to provide more realistic visualizations for both the research team (Fig. 2I) and the surgeon (Fig. 2J). Automating this process (possibly via computer vision grid recognition or and utilizing augmented reality interfaces) could maximize deliberation time and minimize overall surgical time.

In our study, we obtained OFM results through an awake vibrotactile stimulation session. Whether intraoperative stimulation must be performed while the patient is awake is a critical consideration due to the increased time and risk imposed by an awake procedure. Prior studies in humans have demonstrated decreased power in high-gamma frequencies of ECoG recordings under anesthesia.^45^ If accurate delineation of individual finger/fingertip regions can be demonstrated in patients under anesthesia, then the awake portion of the online mapping may not be required.

Additionally, our intraoperative motor mapping did not contribute to motor array placements. Similar to other BMI groups^8,11^ we relied on a priori localization of hand motor areas through preoperative structural and functional imaging of hand knob targets. Final placement was only modified by surface vasculature anatomy (Fig. 3A). It is unclear whether the failure of motor mapping was primarily due to the patient’s inability to fully participate in the motor tasks after awakening, or if motor activity mapped to an ECoG grid could not guide motor anatomy determination beyond structural determination of the hand knob.

As with all surgical procedures, the benefits of functional localization must be weighed against the risks of increased operation time and performing an awake craniotomy. These risks can be minimized by assembling a research team with diverse expertise and experience collaborating together with clinicians skilled in performing awake craniotomies.

## Conclusions

This study demonstrates the potential for online functional mapping, used in conjunction with preoperative imaging and IONM, to improve targeting of finger and fingertip regions of human somatosensory cortex. Accurate targeting of key sensory areas is a necessary step in developing fully dexterous closed-loop neuroprosthesis. Future studies are needed to replicate and improve this approach while demonstrating its utility in a diverse patient population.

## Data Availability

Data is available upon request.

## Acknowledgements

We thank the study participant for his active participation and contributions throughout the study. This research was developed with funding from the Defense Advanced Research Projects Agency’s (DARPA, Arlington) Revolutionizing Prosthetics program (contract number N66001-10-C-4056). The views, opinions, and/or findings expressed are those of the author(s) and should not be interpreted as representing the official views or policies of the Department of Defense or the U.S. Government. Development of experimental setup and support for regulatory submissions associated with this study were provided by a grant from the Alfred E. Mann Foundation. Study software infrastructure and study preparation were developed with internal funding from Johns Hopkins University Applied Physics Laboratory and Johns Hopkins University. Personnel was also supported by NIH (R01NS088606). We thank Dr. Jenessa Malin for her assistance in editing the manuscript.

